# *FMNL2* interacts with cerebrovascular risk factors to alter Alzheimer’s disease risk

**DOI:** 10.1101/2020.08.30.20184879

**Authors:** Neha S. Raghavan, Sanjeev Sariya, Annie J. Lee, Yizhe Gao, Dolly Reyes-Dumeyer, Philip L. De Jager, David A. Bennett, Vilas Menon, Rafael A. Lantigua, Walter A. Kukull, Adam M. Brickman, Jennifer J Manly, Jose Gutierrez, Badri N. Vardarajan, Giuseppe Tosto, Richard Mayeux

**Author notes:** Address Correspondence to: Richard Mayeux, MD, Department of Neurology, Columbia University, 710 West 168th Street, New York, NY 10032.

## Abstract

**INTRODUCTION:** Late-onset Alzheimer’s disease (AD) frequently co-occurs with cerebrovascular disease. We hypothesized that interactions between genes and cerebrovascular risk factors (CVRFs) contribute to AD risk.

**METHODS:** Participants age 65 years or older from five multi-ethnic cohorts (N=14,669) were included in genome-wide association meta-analyses for AD including an interaction factor for a CVRF score created from body mass index, hypertension, heart disease, and diabetes. Significant gene level results were substantiated using neuropathological and gene expression data.

**RESULTS:** At the gene-level, *FMNL2* interacted with the CVRF score to significantly modify AD risk (p= 7.7×10^-7^). A SNP within *FRMD4B*, rs1498837, was nominally significant (p=7.95×10^-7^). Increased *FMNL2* expression was significantly associated with brain infarcts and AD.

**DISCUSSION:** *FMNL2* is highly expressed in the brain and has been associated with ischemic stroke and failures in endosomal trafficking, a major pathway in AD pathology. The results highlight an interaction between *FMNL2* and CVRFs on AD susceptibility.

## 1.0 Introduction

Late-onset Alzheimer’s disease (AD) affects more than 6 million Americans, and is frequently co-morbid with cerebrovascular disease. Neuropathological data indicate that brain tissue from up to 70% of patients with AD have the hallmark neuritic plaques and neurofibrillary tangles with varying degrees of cerebrovascular disease[1-4]. Given the high frequency of this comorbidity, a number of studies have assessed the relationship between cerebrovascular risk factors such as hypertension[5], being overweight[6, 7], diabetes[8, 9] and coronary heart disease[10, 11] and AD. Jagust and colleagues proposed that cerebrovascular disease contributed to AD by perturbing the amyloid-β pathway in addition to causing neurodegeneration[12]. While cerebrovascular risk factors (CVRFs) are associated with AD risk, some reports show no relationship[13], and some show protective effects [13, 14].

The inconsistencies in the relationship between CVRFs and AD could be attributed to age differences, the stage of the disease and medications unaccounted for in these studies. An alternative explanation is that an, as yet unidentified, interaction between genetic variants and CVRFs may affect AD risk. Genetic and genomic studies have indicated a potential relationship between CVRFs and AD. For example, *APOE*, *CLU*, *ABCA7*, and *SORL1*, are part of the lipid metabolism pathway, and provide evidence for a putative molecular relationship to AD. *ABCA7* and *SORL1* are associated with brain infarcts[15]. Using summary statistics from multiple genome-wide associations studies (GWASs), a recent study detected significant pleiotropy between CVRFs and AD[16]. Furthermore, a significant shared genetic contribution to AD and small vessel disease has been reported[17].

Despite putative associations between genes, CVRFs and AD, there has been no genome-wide study of genes or genetic loci that interact with CVRFs to alter AD risk. An interaction would imply that in individuals harboring specific genetic variants, the impact of CVRFs would contribute to AD risk. Genes and loci identified from an interaction analysis may not be found from a typical, main effects GWAS, and would provide novel insights of the effect of cerebrovascular disease on AD risk.

There is evidence that the cumulative burden of multiple CVRFs increases the association with AD risk[18, 19] over investigating single risk factors. Here we use dimensionality reduction of four factors frequently associated with increased risk for cerebrovascular and AD: hypertension, heart disease, diabetes, and body mass index (BMI), to create a CVRF burden score. To augment the sample size, we included data from five different multi-ethnic cohorts facilitating the interaction analyses.

## 2.0 Materials and Methods

### 2.1 Cohorts, Subjects and Study Variables

Participants in the meta-analysis were from the following studies: Washington Heights–Inwood Columbia Aging Project (WHICAP), Estudio Familiar de Influencia Genetica en Alzheimer (EFIGA), the National Alzheimer’s Coordinating Center (NACC), and the Religious Orders Study and Rush Memory and Aging Project (ROSMAP). Details on the cohorts is found in the supplemental methods

We included only participants who were 65 years of age or older, had a clinical assessment of AD and data concerning four CVRFs: any heart disease, hypertension, BMI, and diabetes. Further details of the variables are provided below.

### 2.2 Cerebrovascular Risk Factors Score

Self-reported data from each participant was recorded as a binary indicator (Yes-Ever Had or No-Never Had) for heart disease, hypertension and diabetes. Details on heart conditions varied between groups, so report of any heart disease qualified as presence of heart disease in this study. A quantitative variable was recorded for BMI from the last visit. The PCAMix package[20] computes principal components (PCs) in a mixture of quantitative and qualitative data, and was used to compute PCs from the four cerebrovascular variables. The goal of the dimensionality reduction was to capture the greatest amount of variance accounted for by the four cardiovascular factors, and thus the per participant values from the first principal component from each of the cohorts was used as the individual’s cerebrovascular risk factor score.

### 2.3 Genotyping, Quality Control and Imputation

Genotyping was performed on different platforms separately for each study cohort. Data from all cohorts underwent quality control (QC). Details for QC of each cohort are provided in the supplemental methods.

### 2.4 Statistical Analyses

Study design and analyses are depicted in Supplemental Figure 1. Differences in minor allele frequency across ethnic groups can decrease the power to detect an association, thus we first performed and meta-analyzed gene-based GWAS across the cohorts, followed by SNP-level GWAS. Gene-based tests were performed using the package GESAT[21] and a replication analysis was performed using rareGE[22]. Both packages allow for gene-based analyses with an interaction between environmental variables and multiple variants within a gene using a variance component test; however, different algorithms are used to perform the interaction analysis. We first used GESAT, which permits the use of covariates, allows testing of common variants, and uses ridge regression to estimates effects under the null hypothesis. GESAT and rareGE both output p-values for all tested genes, but do not output a beta value. Therefore, meta-analysis of the results is likely to be inflated because there is no direction associated with the result. To minimize this issue, we chose to only meta-analyze genes that were p<0.01 within cohort and genes with a minimum of five variants. GESAT was performed using default settings. The fixed effect model in rareGE was used to confirm the statistically significant results reported from GESAT. Covariates included in both packages were age, sex, and the first three MDS dimensions to adjust for population substructure and variants with allele frequency greater than 0.01 were included.

SNP based GWAS was performed using R (v 3.5.1) and a logistic model with AD as the outcome. Interaction models were conducted with additive coding plus an interaction factor of SNP*Score. Main effect models and mediation models (including CVRF score as a covariate) were conducted with additive coding for SNPs. For all models used in the NACC cohort, covariates for batch were included as genotyping from ten waves were imputed separately. Covariates in all models were age, sex, and the first three population MDS dimensions.

SNP based tests were meta-analyzed using inverse-weighted methods, and for the gene-based tests were summarized using the sample-size weighted method. Final results are shown for SNPs/genes present in at least 10,000 participants. Significance was set at p = 5×10^-8^ for the genome-wide SNP based tests and p = 2.6×10^-6^ for the gene-based tests after Bonferroni correction for the number of genes tested.

### 2.5 Supportive studies: Analysis plan

We performed supporting studies to further examine our gene of interest from the gene-based GWAS. First, a gene or locus that interacts with cardiovascular or cerebrovascular risk factors likely has some molecular involvement in these vascular pathways, even if indirect. Therefore, we expect an association between the putative gene or locus and cerebrovascular pathology. In order to test this, we first quantified brain infarcts from magnetic resonance imaging (MRI) in WHICAP participants and leveraged pathology data of infarcts from the ROSMAP cohort. We performed association analyses between SNPs within the candidate gene and these cerebrovascular disease measures. Next, we tested whether mRNA expression of the candidate gene from the dorsolateral prefrontal cortex (DLPFC) of ROSMAP participants were associated with brain infarcts. Secondly, we hypothesized that differences in expression of the candidate gene should be associated with AD and AD transcriptional phenotypes. To test this, we performed association analyses of AD diagnosed pathologically and clinically with gene expression described above. In parallel, we performed cell type specific analyses of the expression of our gene of interest.

### 2.6 Measures of cerebrovascular disease

Details of neuropathological evaluation and qualifying determinants of infarcts in the ROSMAP dataset have been reported previously[23]. In the WHICAP cohort, brain infarcts were rated as cavitated lesions seen in axial T1 and T2-weighted FLAIR MRI images. We developed a pathology-informed algorithm to distinguish T1-weighted hypointense lesions larger than 5 mm from dilated perivascular spaces, described previously[24]; details are provided in the supplemental methods. Using R (v 3.5.1), we performed logistic regression for SNPs within our gene of interest from the GWAS with an interaction term of SNP*CVRF score and either the binary outcome variable (present/not present) of: infarcts by MRI in WHICAP or by postmortem examination in ROSMAP as the outcome.

### 2.7 RNA expression analysis

For bulk RNA-sequencing analysis, after rigorous quality control evaluations, we retained RNA-Seq data from the DLPFC of 1092 individuals for downstream analyses; RNA-Seq data were normalized using Trimmed Mean of M-values. Genes with low expression were removed to reduce the influence of technical noise. We assessed the association between the candidate gene expression level and 1) presence of brain infarcts 2) AD diagnosed clinically and pathologically by linear regression. Covariates in the analysis were age, sex, batch effect, postmortem interval, and technical variables and three population structure multidimensional scaling (MDS) dimensions.

### 2.8 Single-nucleus RNA-sequencing analysis

Data was generated from (DLPFC) from the brains of 24 ROSMAP participants[23]. This included 50% women and fell into four categories (six participants per category): (1) Cdx1-pAD0: non-impaired individuals without pathologic criteria for AD by the NIA Reagan scale, (2) Cdx1-pAD1: cognitively non-impaired individuals fulfilling pathologic criteria for AD, (3) Cdx4-pAD0: individuals with dementia who do not fulfill pathologic criteria for AD, and (4) Cdx4-pAD1: individuals with dementia who also fulfilling pathologic criteria for AD. Frozen tissue was thawed and triturated to create a nuclear suspension using the DroNc-Seq protocol [25], and nuclei were loaded onto a 10x Genomics Chromium platform for the generation of cDNA libraries from individual nuclei. Reads were aligned to the GRCh38 genome using CellRanger with default parameters, and with the Ensembl transcriptome annotation (downloaded June 2017, GRCh38.85) modified to include introns for alignment. We recovered a total of ~170,000 post-QC nuclei (with >500 Unique Molecular Identifiers) from the samples, and nuclei were clustered iteratively using the Seurat R package. After clustering, we aggregated the gene expression values for all nuclei belonging to each cell type in each individual.

## 3. Results

### 3.1 Participant characteristics

Data from the five study cohorts included non-Hispanic whites from WHICAP (n=851), NACC (n=8012) and ROSMAP (n=1424), Caribbean Hispanics from the WHICAP and EFIGA study (n=3,404) and African-Americans from the WHICAP study (n=978) (Table 1). Most of the cohort were women and many had a history of hypertension. The ROSMAP cohort was older than the other cohorts.

**Table 1.** Participant demographics included.

|  | NACC | WHICAP<br>White | WHICAP AA | WHICAP<br>CH/EFIGA | ROSMAP |
| --- | --- | --- | --- | --- | --- |
| N | 8012 | 851 | 978 | 3404 | 1424 |
| Ethnicity/Race | NHW | NHW | AA | CH | NHW |
| % Female | 56% | 60% | 72% | 68% | 70% |
| % LOAD | 48% | 21% | 33% | 51% | 34% |
| Mean BMI (s.d.) | 26.24 (4.69) | 26.59 (5.07) | 28.21 (6.17) | 26.89 (5.26) | 26.5 (5.51) |
| % Hypertension | 62% | 66% | 82% | 76% | 62% |
| % Heart Disease | 40% | 44% | 39% | 23% | 22% |
| % Diabetes | 12% | 15% | 29% | 31% | 21% |
| Mean Age (s.d.) | 78.42 (7.69) | 81.01 (7.33) | 80 (7.24) | 75 | 85.93 (6.87) |

### 3.2 Cerebrovascular Risk Factor score

Principal component analysis was performed based on a history of heart disease, hypertension, diabetes, and BMI separately for all cohorts. The squared loadings for principal component 1 for each cohort were as follows: ROSMAP 34.6%, WHICAP White 36.2%, WHICAP African Americans 34.7%, Caribbean Hispanics 38.2%, NACC 35.9% (Supplemental Figure 2). In all cohorts, diabetes and hypertension load most strongly with principal component 1 (Supplemental Figure 2).

### 3.3 Gene-based genome-wide association study

In total, six genes had a p value less than 0.1 in at least four out of the five cohorts, and were tested using GESAT (Table 2). One gene, Formin-like protein 2 (*FMNL2*), met genome-wide significance (p = 7.7×10^-7^). Cohort-level results showed nominal significance (NACC: p = 0.010; African American: p=0.078; WHICAP White: p =0.025; Caribbean Hispanic: p = 0.019; ROSMAP: p= 0.003). This gene by score interaction finding was replicated using rareGE (*FMNL2* p= 0.05).

**Table 2.** Genes analyzed in gene x CVRF score gene-based GWAS.

| Gene | P-value |
| --- | --- |
| *FMNL2* | 7.7 x 10<sup>-7</sup> |
| *NEURL1B* | 6.35 x 10<sup>-5</sup> |
| *HOXB3* | 1.4x10<sup>-4</sup> |
| *CA8* | 1.9 x 10<sup>-4</sup> |
| *HRASLS5* | 5.3 x 10<sup>-4</sup> |
| *C7* | 8.5 x 10<sup>-4</sup> |

### 3.4 Single variant genome-wide association study and mediation analysis

Meta-analysis of single SNP GWAS within the *FMNL2* gene resulted in 85 SNPs with p<0.05 (Supplemental Table 1). Genome-wide, no SNPs significantly interacted with the CVRF score. Manhattan and QQ plots are shown in Figure 1. There was no evidence of inflation in this analysis (λ=1.04). The top signal (p = 7.3910^-07^) was found at rs1498837, located on chromosome 3. In build 38 this SNP is an intronic SNP within *FRMD4B*. The second, strongest independent signal was rs9579210 (p= 2.3×10^-06^), intergenic to *FLT1* and *POMP*. Other top SNPs are reported in Supplemental Table 2.

**Figure 1.**
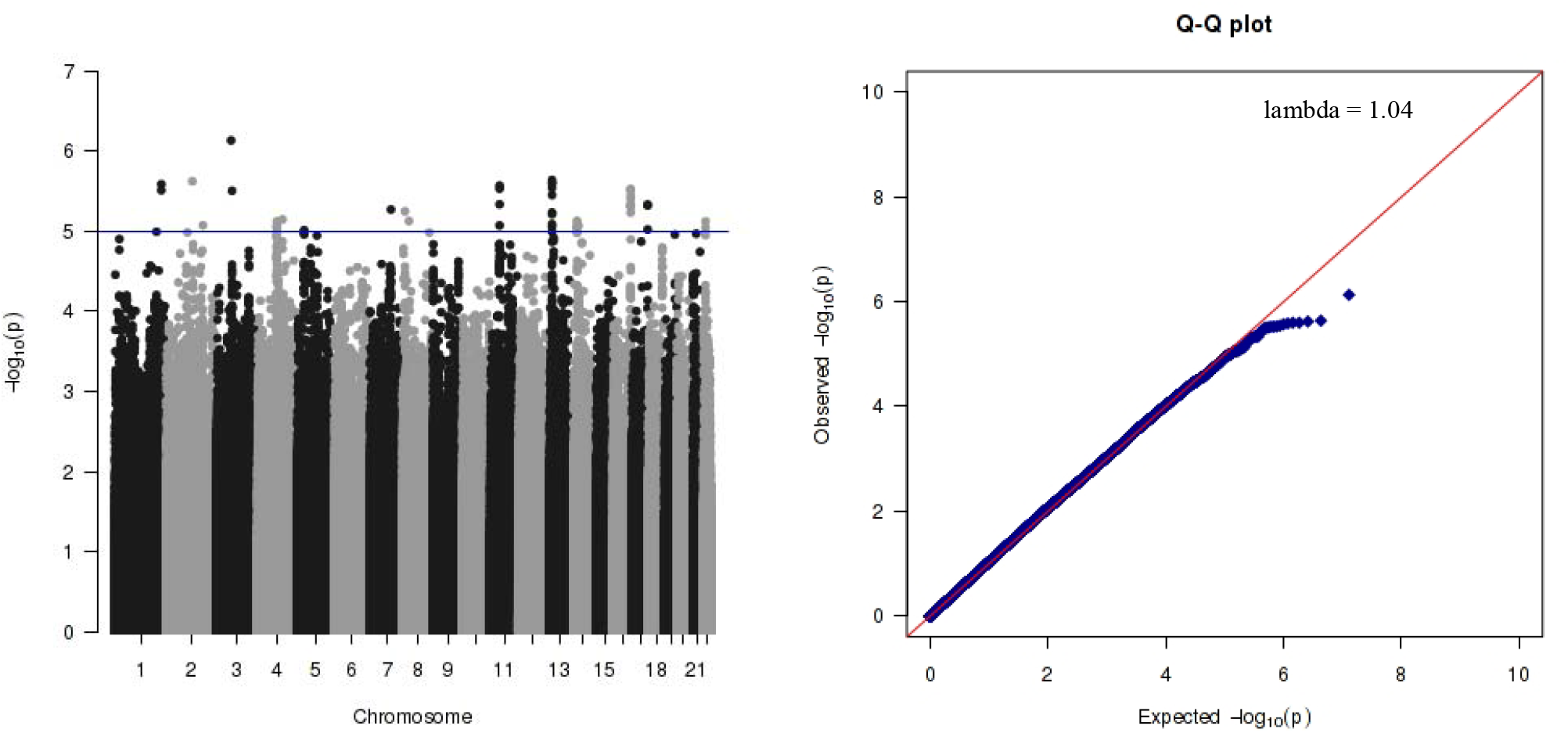
SNP based interaction results; a) Manhattan plot, b) QQ plot

To test for possible mediation effects of the CVRF score, we conducted a main effects model for SNP without CVRF score or any interactions, and subsequently a main effects model including CVRF score as a covariate. Meta-analysis of main effect models in the five cohorts resulted in genome-wide significant results of many SNPs within the *APOE* locus. There was no evidence for inflation in either analysis (Supplemental Figure 3). The top SNP was rs429358 (p =1.54×10^194^) within *APOE*, followed by rs12721051 (p =2.55×10^-165^), an intronic SNP in the *APOC1* gene, within the *APOE* locus. The addition of the CVRF score as a covariate did not identify additional SNPs associated with AD and did not affect the strength of these SNPs (rs429358: p = 4.83×10^-192^; rs12721051: p=2.5×10^-163^). Neither of the two top SNPs from the interaction analysis were nominally significant (p < 0.05) in the main effects model.

### 3.5 Additional genetic and transcriptomic studies

We used three approaches to further investigate the *FMNL2* finding in the gene-based interaction analysis. We assumed that genes interacting with CVRF to modify AD risk might be directly related to molecular pathways in either AD or cerebrovascular disease.

First, we performed SNP-based interaction analyses as before, but constrained to SNPs within *FMNL2*, with measures of cerebrovascular disease as the outcome instead of AD. Outcome data was either presence of brain infarcts measured from MRI from the three ethnic groups that comprise the WHICAP cohort (n = 1,198) or presence of infarcts from neuropathology from the ROSMAP cohort (n = 902). Nine SNPs within *FMNL2* (p<0.05) interacted with CVRF score to affect the risk for brain infarcts, in all three WHICAP cohorts (Supplemental Table 3a). Furthermore, 48 SNPs within *FMNL2* (p<0.05) interacted with CVRF score to affect the risk for infarcts in the ROSMAP neuropathology cohort (Supplemental Table 3b).

Next, we analyzed bulk RNA-sequencing data from the DLPFC in a subset of 1,092 participants from the ROSMAP study. Higher expression of *FMNL2* was associated with greater likelihood of presence of infarcts (p = 0.04). In addition, higher expression of *FMNL2* was associated with greater likelihood of clinically (p = 1.99×10^-8^) and pathologically (p = 0.004) diagnosed AD.

Lastly, we leveraged a set of 24 ROSMAP subjects with single nucleus RNA sequencing data generated from the same brain region to assess the level of *FMNL2* RNA expression in the different subsets of each neocortical cell type. We found that *FMNL2* is expressed in a wide variety of cell types and subtypes (Figure 2); however, it was expressed at a higher concentration in oligodendrocytes and a subset of microglia, similar to another AD susceptibility gene *BIN1*, which is also widely expressed[26] and also implicated in endocytosis. *BIN1* is more highly expressed in oligodendrocytes and microglia (Figure 2). To determine the association between *BIN1* and *FMNL2*, we correlated the mean cell type-specific expression of these two genes across each microglia cluster, corrected for multiple comparisons. In parallel, we plotted the fraction of nuclei and the average z-score of expression for these two genes for each cluster using all 24 subjects. These two analyses together represent cell type-specific expression of these two genes, as well as correlated variation in a given cell type across individuals. The microglia subset 3, which has relatively high expression of both *FMNL2* and *BIN1*, is also increased in frequency in individuals meeting criteria for the pathological diagnosis of AD (Fisher’s exact test p<10^-121^); this enrichment is much higher than for microglial subset 0 (p<10^-32^) and the other microglial subsets (subsets 1,2,4, and 5) which are not altered at all in AD (Cain A, et al; personal communication). In microglia subtype 3, the expression of *BIN1* and *FMNL2* are correlated (Pearson’s r=0.14, p=0.01); however, no such correlation is seen when evaluating all microglia together or in any of the other three subtypes of microglia (Figure 2).

**Figure 2.**
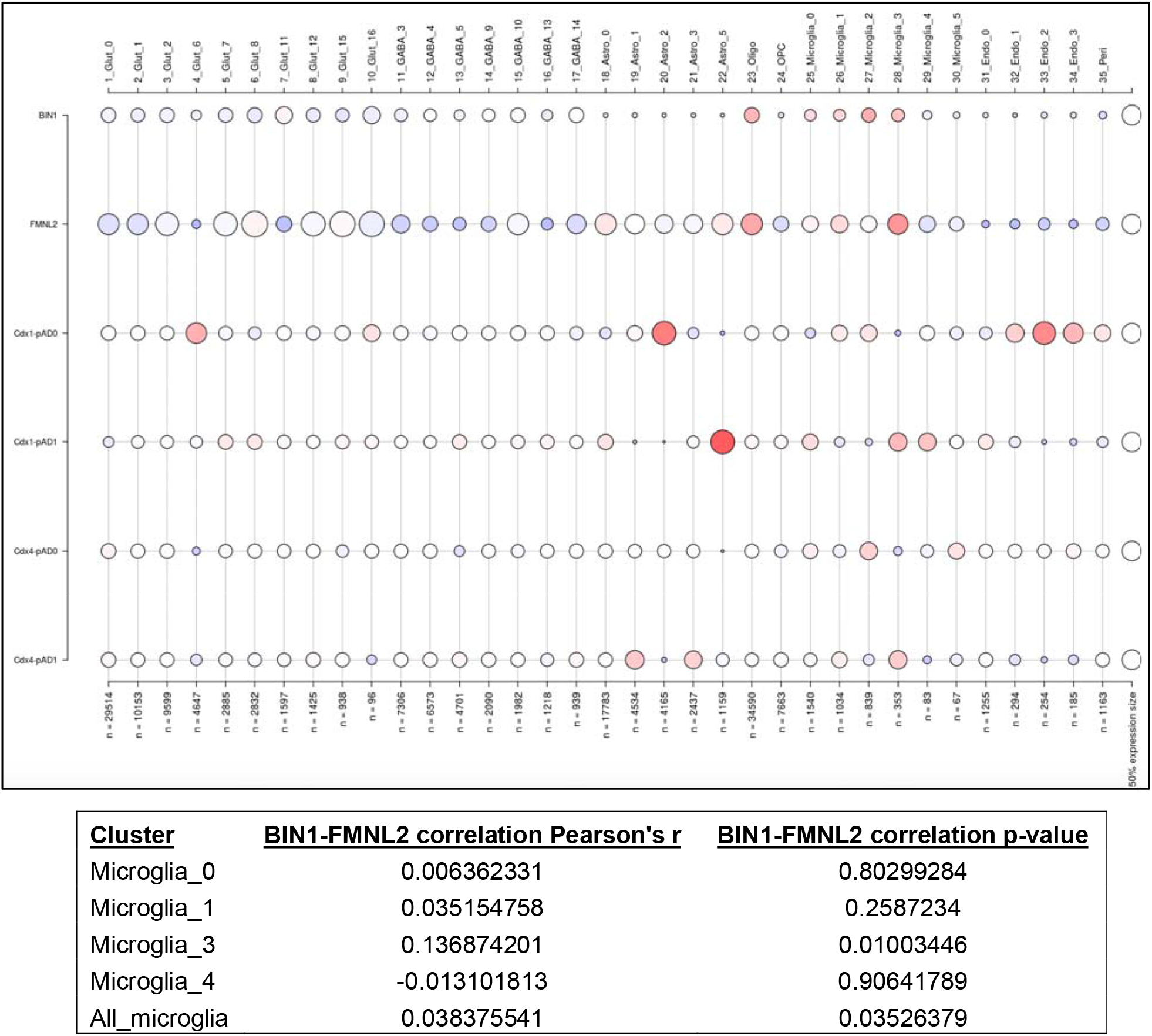
Single cell expression from ROSMAP dorsolateral frontal cortex. Expression levels of genes and participant diagnosis categories within cell type specific clusters. Cell type specific clusters: Cdx1-pAD0: non-impaired individuals without pathologic criteria for AD by the NIA Reagan scale, (2) Cdx1-pAD1: cognitively non-impaired individuals fulfilling pathologic criteria ia for AD, (3) Cdx4-pAD0: individuals with dementia who do not fulfill pathologic criteria for AD, D, and (4) Cdx4-pAD1: individuals with dementia who also fulfilling pathologic criteria for AD.

## 4. Discussion

A major effort to identify genes and loci associated with AD risk has been underway for nearly a decade[27, 28]. Many of these genes have been replicated and confirmed in other studies[29, 30]. The focus now is to understand the mechanisms by which variants in genes lead to disease. Epidemiological and neuropathological research has established an unmistakable, intertwined relationship between AD and cerebrovascular disease. Yet, our understanding of the course of this inter-dependent relationship is unclear. To further probe the relationship of cerebrovascular disease with AD based on genotype, we used data from five multi-ethnic cohorts to perform an AD genome-wide association study of the interaction effects of CVRFs and genetic susceptibility. We found that *FMNL2*, formin-like 2, significantly interacted with a CVRF score among individuals with clinical and pathological AD. We also found that the association between CVRF score and AD was significantly different based on SNP genotypes within *FMNL2* and brain gene expression of *FMNL2* was associated with increased risk of brain infarcts and AD. Furthermore, our results indicate that *FMNL2* is highly expressed in the microglia and correlates with *BIN1* expression.

Interaction effects may explain why, aside from APOE, the large majority of other genes significantly associated with AD have a very modest effect size. Furthermore, genetic studies may obscure CVRF association studies of AD, contributing to some of the conflicting results in the literature. Previous studies of gene-environment interactions in AD have focused on candidate genes, stemming from prior AD or vascular disease GWASs (e.g.[31, 32]). We focused on a genome-wide, unbiased interaction approach to identify novel genes that interact with CVRFs to increase susceptibility to AD.

A strength of the current study is the inclusion of three racial/ethnic groups, which is particularly important in the study of genetics and cerebrovascular disease, for two reasons. First, specific variants associated with disease within a gene can differ between ethnic groups based on linkage disequilibrium (LD) structure; identifying significant SNPs through a meta-analysis across cohorts and collapsing SNPs within gene for gene-based analyses increases the generalizability of the results. Secondly, there are differences in rates of cerebrovascular disease and associated risk factors between non-Hispanic Whites, African Americans, and Caribbean Hispanics. Clinically, these results could have important implications for personalized treatment plans, suggesting more aggressive treatments of cardiovascular and cerebrovascular risk factors in persons with specified genotypes.

*FMNL2*, encodes a formin-related protein. Formins are important in regulating actin and microtubules, with cellular effects and consequences in cell morphology and cytoskeletal organization. Silencing of FMNL2 results in decreased efficiency of cell migration[33]. *FMNL2* is most highly expressed in the brain and has been related to cardiovascular and cerebrovascular disease. For example, in a small GWAS[34] (n = 1,816) of ischemic stroke in young individuals (15-49), a SNP within *FMNL2* was the most significant finding (rs2304556 P = 1.18×10-7), although it did not reach statistical significance. Interestingly, we found that increased *FMNL2* expression was associated with brain infarcts and AD independently, suggesting that this gene may be involved in a shared molecular mechanism with cerebrovascular disease. Knock-down of FMNL2 and related FMNL3 protein was found to result in the defective sorting into late endosomes and lysosomes[35]. Abnormalities in the endosomal-lysosomal network are known to be important to the mechanism of neurodegeneration and AD[36].

While there were no genome-wide significant findings in the SNP based analyses, there were suggestive findings that we highlight here. The most significant suggestive SNP was within the gene *FRMD4B*, which is also highly expressed in the brain. An intronic SNP (rs6787362) within *FRMD4B* has been associated with ischemic heart failure[37, 38], atrial fibrillation[39],and diastolic blood pressure[40]. The other top suggestive SNP is intergenic to POMP and FTL1, which encodes a member of the vascular endothelial growth factor receptor (VEGFR) and is important in angiogenesis and vasculogenesis.

Our findings infer that the involvement of genes in cerebrovascular pathways may predispose individuals to AD. A lack of association of vascular-related genes such as *APOE* may indicate that the association of these genes is stable across those with and without cardiovascular risk factors present. Thus, the most parsimonious conclusion of this study is that when the cerebrovascular burden, both genetic and phenotypic, is increased, AD risk follows.

While this study has a number of strengths, we recognize that there are also a number of limitations in this study. First, we relied on self-report of CVRFs, possibly missing individuals who report incorrectly. Secondly, we did not account for possible effects of medications used to treat cardiovascular disease. However, both effects would be non-differential. Furthermore, we could not be more granular with some cardiovascular or cerebrovascular diagnoses as some cohorts for example only reported “any heart disease,” while others included information on the specific heart condition. Therefore, both mechanical and acquired heart disease are considered equally. Finally, we believe that a larger sample size will be necessary to identify individual SNPs and additional genes that significantly interact with cerebrovascular disease risk factors.

Taken together this report has implicated interactions between a gene, *FMNL2* and the presence of CVRFs to affect AD risk. These results indicate that the clinical risk for developing AD in individuals in the presence of cerebrovascular risk factors may differ by genetic profiles. Specifically, presence of risk factors and variants in genes involved in cerebrovascular disease may have multiplicative effects on disease risk. Further work expanding on the relationship of genes and loci identified here, along with expanded basic molecular work will move us closer to conceptualizing the relationship between AD and cerebrovascular disease.

## Data Availability

All the data included in the paper can be accessed after request.
Genetic and phenotype data for WHICAP can be requested:https://cumc.co1.qualtrics.com/jfe/form/SV_6x5rRy14B6vpoqN
Genetic and phenotype data for EFIGA can be requested: https://cumc.co1.qualtrics.com/jfe/form/SV_dmck0uV3A91pmzb
Genetic and phenotype data for ROSMAP can be requested: https://www.radc.rush.edu/
Genetic and phenotype data for NACC can be requested:https://www.alz.washington.edu/WEB/landingRequest.html, https://www.niagads.org/user/login?destination=data/request/new_request/

https://cumc.co1.qualtrics.com/jfe/form/SV_dmck0uV3A91pmzb

https://www.radc.rush.edu/

https://www.alz.washington.edu/WEB/landingRequest.html

https://www.niagads.org/user/login?destination=data/request/new_request/

## Acknowledgements

This work was supported by funding from the National Institute on Aging (NIA), by the National Institutes of Health (NIH) (1RF1AG054023, 5R37AG015473, RF1AG015473, R56AG051876), and the National Center for Advancing Translational Sciences, NIH through Grant Number UL1TR001873 and TL1TR001875. The National Alzheimer’s Coordinating Center is supported by U01AG016976. ROSMAP was supported by NIA grants P30AG10161, R01AG15819, R01AG17917, U01AG46152, U01AG61356.

## References

[1] Attems J, Jellinger KA. The overlap between vascular disease and Alzheimer’s disease--lessons from pathology. BMC Med. 2014;12:206.

[2] Brenowitz WD, Keene CD, Hawes SE, Hubbard RA, Longstreth WT, Jr., Woltjer RL, et al. Alzheimer’s disease neuropathologic change, Lewy body disease, and vascular brain injury in clinic- and community-based samples. Neurobiol Aging. 2017;53:83–92.

[3] Kapasi A, DeCarli C, Schneider JA. Impact of multiple pathologies on the threshold for clinically overt dementia. Acta Neuropathol. 2017;134:171–86.

[4] Schneider JA, Arvanitakis Z, Bang W, Bennett DA. Mixed brain pathologies account for most dementia cases in community-dwelling older persons. Neurology. 2007;69:2197–204.

[5] Skoog I, Lernfelt B, Landahl S, Palmertz B, Andreasson LA, Nilsson L, et al. 15-year longitudinal study of blood pressure and dementia. Lancet. 1996;347:1141–5.

[6] Gustafson D, Rothenberg E, Blennow K, Steen B, Skoog I. An 18-year follow-up of overweight and risk of Alzheimer disease. Arch Intern Med. 2003;163:1524–8.

[7] Luchsinger JA, Cheng D, Tang MX, Schupf N, Mayeux R. Central obesity in the elderly is related to late-onset Alzheimer disease. Alzheimer Dis Assoc Disord. 2012;26:101–5.

[8] Cheng D, Noble J, Tang MX, Schupf N, Mayeux R, Luchsinger JA. Type 2 diabetes and late-onset Alzheimer’s disease. Dement Geriatr Cogn Disord. 2011;31:424–30.

[9] Luchsinger JA, Tang MX, Stern Y, Shea S, Mayeux R. Diabetes mellitus and risk of Alzheimer’s disease and dementia with stroke in a multiethnic cohort. Am J Epidemiol. 2001;154:635–41.

[10] Justin BN, Turek M, Hakim AM. Heart disease as a risk factor for dementia. Clin Epidemiol. 2013;5:135–45.

[11] Ott A, Breteler MM, de Bruyne MC, van Harskamp F, Grobbee DE, Hofman A. Atrial fibrillation and dementia in a population-based study. The Rotterdam Study. Stroke. 1997;28:316–21.

[12] Jagust W. Imaging the evolution and pathophysiology of Alzheimer disease. Nat Rev Neurosci. 2018;19:687–700.

[13] Tosto G, Bird TD, Bennett DA, Boeve BF, Brickman AM, Cruchaga C, et al. The Role of Cardiovascular Risk Factors and Stroke in Familial Alzheimer Disease. JAMA Neurol. 2016;73:1231–7.

[14] Nourhashemi F, Deschamps V, Larrieu S, Letenneur L, Dartigues JF, Barberger-Gateau P, et al. Body mass index and incidence of dementia: the PAQUID study. Neurology. 2003;60:117–9.

[15] Farfel JM, Yu L, Buchman AS, Schneider JA, De Jager PL, Bennett DA. Relation of genomic variants for Alzheimer disease dementia to common neuropathologies. Neurology. 2016;87:489–96.

[16] Broce IJ, Tan CH, Fan CC, Jansen I, Savage JE, Witoelar A, et al. Dissecting the genetic relationship between cardiovascular risk factors and Alzheimer’s disease. Acta Neuropathol. 2019;137:209–26.

[17] Traylor M, Adib-Samii P, Harold D, Alzheimer’s Disease Neuroimaging I, International Stroke Genetics Consortium UKYLSDNAr, Dichgans M, et al. Shared genetic contribution to Ischaemic Stroke and Alzheimer’s Disease. Ann Neurol. 2016;79:739–47.

[18] Luchsinger JA, Reitz C, Honig LS, Tang MX, Shea S, Mayeux R. Aggregation of vascular risk factors and risk of incident Alzheimer disease. Neurology. 2005;65:545–51.

[19] Reitz C, Tang MX, Schupf N, Manly JJ, Mayeux R, Luchsinger JA. A summary risk score for the prediction of Alzheimer disease in elderly persons. Arch Neurol. 2010;67:835–41.

[20] Chavent M, Kuentz, V., Labenne, A., Liquet, B., Saracco, J. PCAmixdata: Multivariate Analysis of Mixed Data. R package version 3.1. 2017.

[21] Lin X, Lee S, Christiani DC, Lin X. Test for interactions between a genetic marker set and environment in generalized linear models. Biostatistics. 2013;14:667–81.

[22] Chen H, Meigs JB, Dupuis J. Incorporating gene-environment interaction in testing for association with rare genetic variants. Hum Hered. 2014;78:81–90.

[23] Schneider JA, Bienias JL, Wilson RS, Berry-Kravis E, Evans DA, Bennett DA. The apolipoprotein E epsilon4 allele increases the odds of chronic cerebral infarction [corrected] detected at autopsy in older persons. Stroke. 2005;36:954–9.

[24] Gutierrez J, Elkind MS, Cheung K, Rundek T, Sacco RL, Wright CB. Pulsatile and steady components of blood pressure and subclinical cerebrovascular disease: the Northern Manhattan Study. Journal of hypertension. 2015.

[25] Habib N, Avraham-Davidi I, Basu A, Burks T, Shekhar K, Hofree M, et al. Massively parallel single-nucleus RNA-seq with DroNc-seq. Nat Methods. 2017;14:955–8.

[26] Taga M, Petyuk VA, White C, Marsh G, Ma Y, Klein HU, et al. BIN1 protein isoforms are differentially expressed in astrocytes, neurons, and microglia: neuronal and astrocyte BIN1 are implicated in tau pathology. Mol Neurodegener. 2020;15:44.

[27] Lambert JC, Grenier-Boley B, Harold D, Zelenika D, Chouraki V, Kamatani Y, et al. Genome-wide haplotype association study identifies the FRMD4A gene as a risk locus for Alzheimer’s disease. Mol Psychiatry. 2013;18:461–70.

[28] Jun G, Ibrahim-Verbaas CA, Vronskaya M, Lambert JC, Chung J, Naj AC, et al. A novel Alzheimer disease locus located near the gene encoding tau protein. Mol Psychiatry. 2016;21:108–17.

[29] Kunkle BW, Grenier-Boley B, Sims R, Bis JC, Damotte V, Naj AC, et al. Genetic meta-analysis of diagnosed Alzheimer’s disease identifies new risk loci and implicates Abeta, tau, immunity and lipid processing. Nature genetics. 2019;51:414–30.

[30] Bis JC, Jian X, Kunkle BW, Chen Y, Hamilton-Nelson KL, Bush WS, et al. Whole exome sequencing study identifies novel rare and common Alzheimer’s-Associated variants involved in immune response and transcriptional regulation. Mol Psychiatry. 2018.

[31] Cambronero FE, Liu D, Neal JE, Moore EE, Gifford KA, Terry JG, et al. APOE genotype modifies the association between central arterial stiffening and cognition in older adults. Neurobiol Aging. 2018;67:120–7.

[32] Weng PH, Chen JH, Chen TF, Sun Y, Wen LL, Yip PK, et al. CHRNA7 Polymorphisms and Dementia Risk: Interactions with Apolipoprotein epsilon4 and Cigarette Smoking. Sci Rep. 2016;6:27231.

[33] Block J, Breitsprecher D, Kuhn S, Winterhoff M, Kage F, Geffers R, et al. FMNL2 drives actin-based protrusion and migration downstream of Cdc42. Curr Biol. 2012;22:1005–12.

[34] Cole JW, Stine OC, Liu X, Pratap A, Cheng Y, Tallon LJ, et al. Rare variants in ischemic stroke: an exome pilot study. PLoS One. 2012;7:e35591.

[35] Kage F, Steffen A, Ellinger A, Ranftler C, Gehre C, Brakebusch C, et al. FMNL2 and −3 regulate Golgi architecture and anterograde transport downstream of Cdc42. Sci Rep. 2017;7:9791.

[36] Nixon RA, Yang DS. Autophagy failure in Alzheimer’s disease--locating the primary defect. Neurobiol Dis. 2011;43:38–45.

[37] Cappola TP, Li M, He J, Ky B, Gilmore J, Qu L, et al. Common variants in HSPB7 and FRMD4B associated with advanced heart failure. Circ Cardiovasc Genet. 2010;3:147–54.

[38] Matkovich SJ, Van Booven DJ, Cappola TP, Dorn GW, 2nd. Association of an intronic, but not any exonic, FRMD4B sequence variant and heart failure. Clin Transl Sci. 2010;3:134–9.

[39] Nielsen JB, Thorolfsdottir RB, Fritsche LG, Zhou W, Skov MW, Graham SE, et al. Biobank-driven genomic discovery yields new insight into atrial fibrillation biology. Nature genetics. 2018;50:1234–9.

[40] Houwing-Duistermaat JJ, Helmer Q, Balliu B, van den Akker E, Tsonaka R, Uh HW. Gene analysis for longitudinal family data using random-effects models. BMC Proc. 2014;8:S88.

